# The Impact of Climate Change on the Prevalence of Mental Illness Symptoms

**DOI:** 10.1101/2021.08.06.21261722

**Authors:** Molly Monsour, Emily Clarke-Rubright, Wil Lieberman-Cribbin, Christopher Timmins, Emanuela Taioli, Rebecca M. Schwartz, Samantha S. Corley, Anna M. Laucis, Rajendra A. Morey

## Abstract

**Background:** The repercussions of climate change threaten the population with an increased prevalence of extreme climate events. We explored the impact of climate change induced sea level rise (SLR) and tropical cyclone (TC) exposure on mental illness symptom prevalence.

**Methods:** Using three datasets, TC exposure scores were calculated for each subject to determine how exposure affects posttraumatic stress disorder (PTSD), anxiety, and major depressive disorder (MDD) symptom prevalence. Inundation mapping of various SLR and storm surge (SS) scenarios were performed for the susceptible region of Miami-Dade and Broward counties to determine the population impact of flooding.

**Results:** We found an elevated risk of mental illness symptoms from exposure to more high-intensity TCs and identified demographic variables that may contribute to this risk. Furthermore, inundation mapping demonstrated severe and widespread impact of SLR and SS on the mental health of communities.

**Limitations:** This study did not include data directly measuring comorbidity, resilience, preparedness, or ability to adapt to climate change. Also, multiple imputation using chained equations may have been imperfect. Finally, when conducting inundation mapping, static mapping may overestimate flooding severity.

**Conclusion:** The impacts of climate change have been frequently studied in terms of physical health, natural disaster prevalence, and economic impacts, but rarely on mental health burden. However, it is vital that national, state, and local governments develop and deploy plans to address mental health needs along with expenditures for protecting infrastructure, the economy, and physical health from the combined effects of SLR and climate change-induced natural disasters.

## INTRODUCTION

With projected drastic increases in global temperature (Watts et al., 2019), the population will undeniably suffer adverse consequences, such as heatwaves, floods, droughts (Berry et al., 2018), inadequate nutrition, polluted air (Trombley et al., 2017), and new communicable diseases (Bourque and Willox, 2014). The grave impacts of climate change have been frequently studied in terms of physical health, natural disaster prevalence, and environmental changes (Bourque and Willox, 2014); in comparison, however, literature on the relationship between climate change and mental health is less available (Berry et al., 2010).

As the world faces the detrimental consequences of climate change, the impact on individuals’ mental health is important to consider. Higher temperatures have been associated with mental health decline (Obradovich et al., 2018), increased aggression, violent crime (Anderson, 2001), and increased prevalence of suicide (Maes et al., 1994). The increase in natural disasters due to climate change may also lead to increased rates of posttraumatic stress disorder (PTSD), anxiety disorders, and major depressive disorder (MDD) (Makwana, 2019; Schwartz et al., 2017b; Schwartz et al., 2017c; Schwartz et al., 2016; Schwartz et al., 2015; Schwartz et al., 2018). Given the potential effects of climate change on mental health, research into the psychological and brain health impacts of global warming are imperative.

### Sea Level Rise (SLR) and Storm Surge

SLR-induced flooding due to climate change poses a threat to coastal cities (Sweet and Park, 2014). Rapidly rising sea levels produce thermal expansion of ocean surface waters, melting glaciers, elevated evaporation and precipitation, and the slowing of the Atlantic Meridional Overturning Circulation, all of which result from global warming (Hoegh-Guldberg et al., 2018; Wdowinski et al., 2015; Williams et al., 2008). Various projections for 2100 predict 0.2 – 2.0 meters SLR, depending on global efforts to reduce the impacts of climate change (IPCC, 2013; NOAA, 2012; Strauss et al., 2014). As sea levels rise, flooding will occur more frequently and more severely impact coastal areas (Sweet and Park, 2014). SLR is a rapidly approaching crisis for which communities must prepare.

In addition, storm surge (SS), defined as increased sea levels due to low atmospheric pressure and/or strong winds (Pachauri and Reisinger, 2007), caused by tropical cyclones (TCs) will further increase flood risk in coastal communities (Dasgupta et al., 2010; Galea et al., 2007; Mcbride et al., 2006; Pielke et al., 2005; Southeast Florida Regional Climate Change Compact Sea Level Rise Work Group, 2015). TC incidence and SS may increase as carbon emissions allow sea surface temperatures to reach the 26.5°C threshold for TC development (Goldenberg et al., 2001; Gray, 1968). Some reports, however, suggest that increases in CO_2_ and temperature will lower overall cyclone frequency by 5-30% but increase the number of category 4 and 5 TCs by up to 25% (Walsh et al., 2016). Category 4 and 5 TCs correspond to a 5-20% increase in rainfall and extreme SS, further contributing to flood risk (National Park Service, 2019). The distressing impacts of climate change induced natural disasters will be felt globally, but coastal areas such as South Florida are especially vulnerable.

### Region of Interest

By 2100, SLR is predicted to impact up to 43 million people (Hauer, 2020). Of these millions, half are from Florida, and 25% are Miami residents (Nicholls et al., 2011). The rate of SLR in southeast Florida is almost three times as rapid as global SLR (Wdowinski et al., 2015) due to its minimally sloped topography, low elevation, variable salinity, non-uniform ocean warming, ocean circulation, and deglaciation (Hay, 2015; Milne, 2009; Stammer, 2008; Wunsch, 2007; Zhang, 2010). According to Esri projections, Miami-Dade and Broward counties have the second and third highest TC occurrence in the country, with a history of fourteen and twelve major (category 3-5) TCs, respectively (esri). If inundation levels reach 1 meter in Florida, 2,555 miles of road, 35 public schools, $145 billion worth of property, and 300,000 homes would be affected (Strauss et al., 2014). Despite this high risk of damage from SLR, only 2% of Florida, 4% of Miami Dade, and 4% of Broward county have preparation plans for this type of devastation (Strauss et al., 2014). Considering these risk factors, it is pertinent to study the potential risks of SLR and SS in these counties.

### Mental Health, Flooding, and TCs

With these grave repercussions of climate change and research supporting that natural disasters increase the risk of PTSD, anxiety, and MDD diagnoses, it is vital that researchers investigate the mental health impacts of flooding and TCs (Lieberman-Cribbin et al., 2017; Makwana, 2019). After the Great Midwest Floods of 1993, the 1997 flood in Central Valley, CA, the Iowa floods, the Kentucky floods of 1981 and 1984, and the 2013-2014 floods in England, the prevalence of mental illness was elevated (Ginexi et al., 2000; Jermacane et al., 2018; McMillen et al., 2002; Phifer et al., 1988; Waelde et al., 2000; Waite et al., 2017). Similarly, elevated rates of mental illness were reported after TCs Katrina, Maria, Ike, the 2004 Florida TCs, Sidr, and Orissa, (Acierno et al., 2007; An et al., 2019; Ferré et al., 2018; Galea et al., 2007; Kar, 2004; Lowe et al., 2013; Nahar et al., 2014; Pietrzak et al., 2012; Turpin). These studies verify the strong link between climate change-induced flooding, TC incidence, and mental health.

The present study used previously collected data from the survivors of TC Ike, the 2004 Florida TCs, and TC Sandy which examined post-disaster population-level prevalence of mental illness diagnoses or symptoms. This data was used to create projection models for prevalence of PTSD, anxiety, or MDD symptoms resulting from climate change-induced TC exposure. Using digital elevation models (DEM) of Miami-Dade and Broward counties, we mapped the percentage of inundated land in relation to SS height and SLR to determine the percentage of the population impacted. Our research will assist policy makers in local, state, and national governments and nongovernmental organizations to better understand, plan, and prepare for the detrimental impacts of climate change on mental illness symptoms.

## METHODS

The methods consist of four main parts. First, datasets from various sites were harmonized and organized to build logistic regression models relating various demographics, previous trauma, post-disaster support, and TC exposure to the elevated risk of PTSD, anxiety, or MDD symptoms. For more details about the data cleaning and variable definitions, refer to Online Resource 3. Secondly, further analysis of the data considered how TC exposure alone contributes to the elevated risk of mental illness symptoms. Inundation mapping was then performed on Miami-Dade and Broward Counties to determine the percentage of the population that will be impacted by various SLR and SS scenarios. Finally, results from the inundation mapping and logistic regression models were used to project the expected number of excess cases of mental illness symptoms in Miami-Dade and Broward counties.

### Projection Models

Using the R function *glm* (MASS package), logistic regression models were built based on three previously collected datasets from TC Ike, TC Sandy, and the 2004 Florida TCs (Acierno et al., 2007; National Center for Disaster Mental Health Research et al., 2016; Schwartz et al., 2017a; Venables and Ripley, 2002). Information on data organization and harmonization and a detailed explanation for calculating the TC Exposure Score per site can be found in the Online Resources (OR 1-3). To avoid bias due to differences in missing data across sites, the data was imputed in R using multiple imputation by chained equations via the *mice* function (van Buuren and Groothuis-Oudshoorn, 2011). Multiple imputation uses the value of many correlated variables to impute the value of a missing variable in individual subjects. Prior to imputation, *scale* was used to center the health score, income, emotional support, informational support, number of previous traumas, and number of days displaced by subtracting the column mean (omitting NAs) and then dividing the centered columns by the standard deviation (R Core Team, 2020). Using *mice*, the predictor matrix and methods of imputation were determined. Incidence of PTSD, anxiety, or MDD symptoms after TC exposure were set to 0 in the matrix. Because TC Exposure was calculated differently for each site, the TC score was also set to 0 in the matrix. Using the standard method, predictive mean matching, and the previously outputted predictor matrix, another imputation was performed with *mice*. The *complete* function (Wickham and Henry, 2020) was then used to fill in the missing data. Once the missing data points were imputed, we selected variables from the new datasets which we hypothesized would impact the risk of mental illness to develop our models with *glm* (Venables and Ripley, 2002). Using these selected variables as covariates, the *glm* output included the β values for demographic variables, previous trauma exposure, post-disaster support, and normalized TC exposure as related to elevated risk of PTSD, anxiety, or MDD symptoms. The *step* function was used to further simplify the logistic regression results (R Core Team, 2020) (OR 2; Fig. 1). Another model was created to determine the effect size of TC exposure on PTSD, anxiety, or MDD symptoms risk without other variables (OR 2; Fig. 1). Using these effect sizes, *ggplot* (Wickham, 2016) was used to graph the elevated risk of PTSD, anxiety, or depression symptoms versus normalized TC exposure scores per site (Fig. 3). The *meanz* function (metap package) (Dewey, 2020) was used to combine the p-values for the effects of TC exposure on PTSD, anxiety, or depression symptom risk as specified by the mean of z method (Becker, 1994) (Fig. 1).

**Fig. 1.**
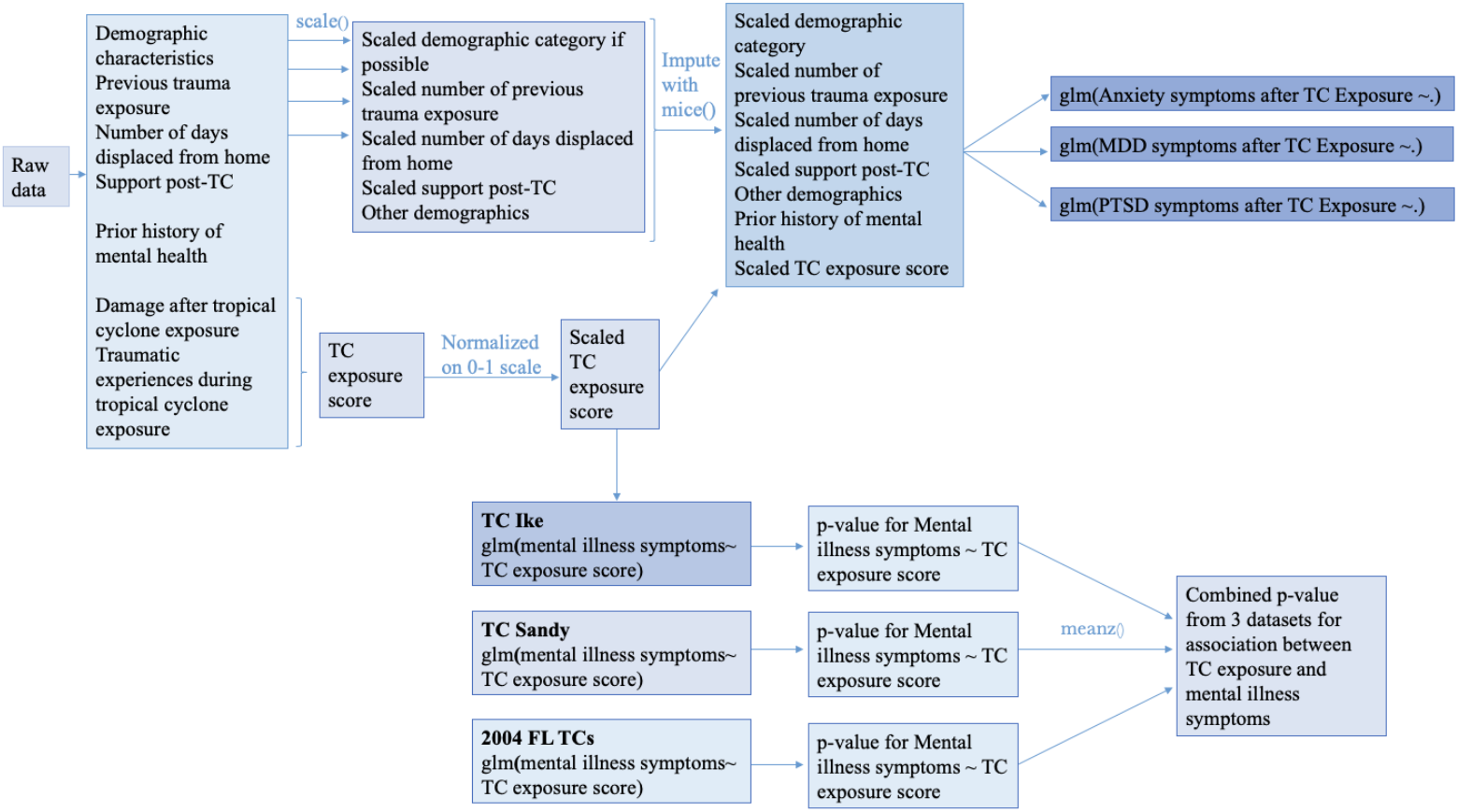
General regression model development in R (TC = Tropical Cyclone)

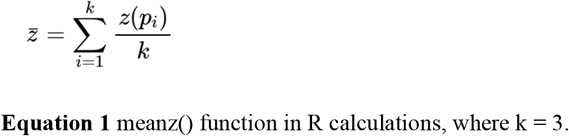

### Mapping inundation in ArcGIS

Static inundation mapping was used to analyze the impact of various SLR scenarios for Miami Dade and Broward counties (Fig. 2). Static mapping assumes that land below a specified elevation will experience inundation (Gallien, 2016). Based on current projections, the SLR scenarios mapped were 0.5, 1.0, 1.5, and 2.0 meters (Harris, 2017; IPCC, 2013; NOAA, 2012; Strauss et al., 2014; USACE U.S. Army Corps of Engineers, 2014). We used 1/3 arc-second DEMs as an elevation data source (U.S. Geological Survey, 2019). While some research on SLR inundation will use MHHW as a reference water level to reflect high tides, the difference between MHHW and North American Vertical Datum of 1988 (NAVD88) for the Virginia Key tidal station (representative of Miami-Dade county’s tides) is only 0.062 m. For the South Port Everglades tidal station (representative of Broward county’s tides), the difference is only 0.163 meters (NOAA, 2018; Zhang, 2010). Thus, inundation levels were relative to the NAVD88 and the maps referenced the North American Datum of 1983 (NAD83). To determine inundation levels, the ArcGIS raster calculator tool marked cells below a certain elevation as inundated. This raster was transformed to polygons using the raster to polygon tool. By clipping the population data polygons from the ArcGIS layer, ACS Population Variables (U.S. Census Bureau, 2019) to the inundation polygons, the number of people impacted by the flooding was calculated with equation 2 (Fig 2; OR 4).

**Fig. 2.**
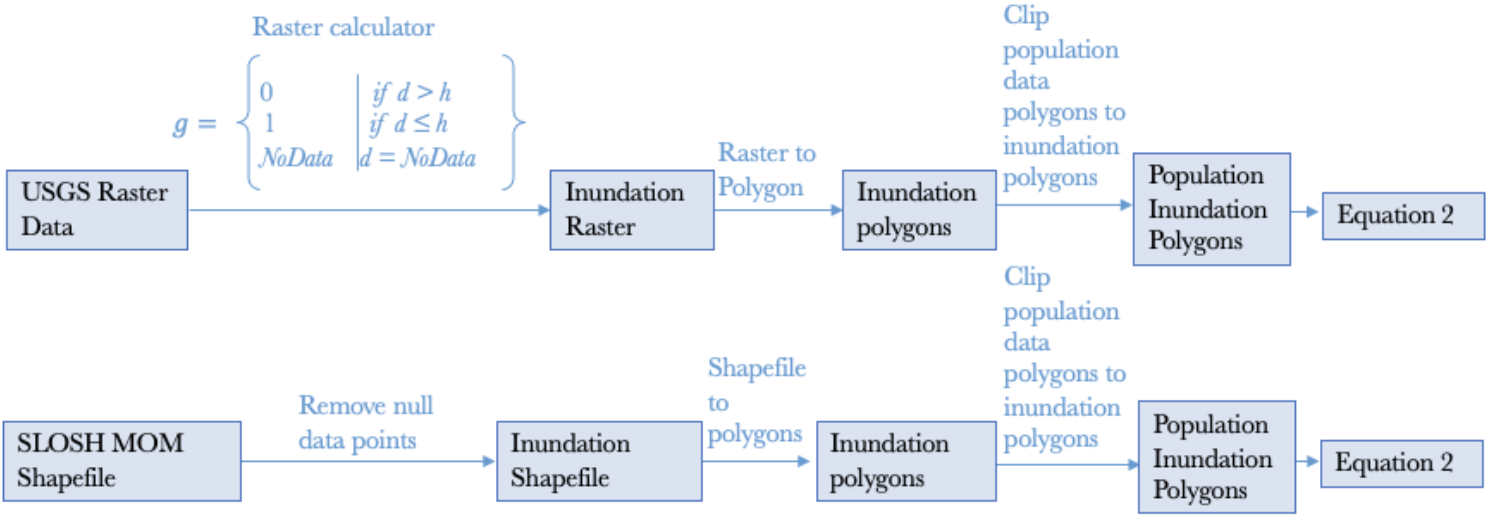
Inundation mapping in ArcGIS

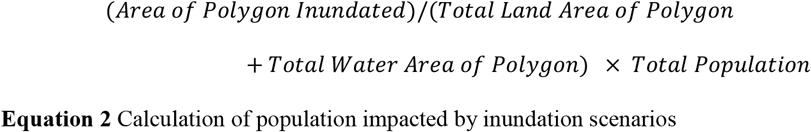

To map various SS scenarios, the National Weather Service’s computerized model, Sea, Lake, and Overland Surge from TCs (SLOSH) (NWS, 2011) was used. The SLOSH software considers the pressure, radius of maximum winds, location, direction, and forward speed of a TC, in addition to the area of interest’s topography and bathymetry to calculate SS height. SLOSH creates multiple Maximum Envelope of Water (MEOW) scenarios for various TC conditions. Taking the maximum SS height (relative to high-tide sea levels) for all TCs of a given category, the Maximum of MEOW (MOM) can be developed for various regions. For this study, MOMs for category 1-5 TCs were downloaded as shapefiles, projected onto the NAD83, and cleaned to remove null data points. The MOMs were then clipped relative to the population data.

While these SS scenarios result in extreme inundation, they are relative to current sea levels and fail to reflect the SLR noted above. For the combined SLR and SS scenarios, the Saffir Simpson TC Scale’s predictions for SS heights were added to the SLR scenario water levels for static inundation mapping values.

### Projected Prevalence of Mental Illness Symptoms from SLR and TC Intensity Levels

Relating the inundation mapping and logistic regression models, we predicted the number of cases of symptoms of anxiety disorder, MDD, or PTSD in Miami-Dade and Broward counties (OR 9). The datasets used did not include flood levels experienced by each subject. Thus, our methods for calculating TC exposure score do not directly correlate to flood heights (OR 3). Rather, we designated a scaled TC Exposure Score of 1, which is the most intense exposure, to be the most devastating SLR + SS inundation scenario (2.0 m SLR and category 5 TC). In this scenario, 4,442,607 people would be affected by flooding. For each site and each mental illness, we multiplied the predicted elevated risk of symptoms given a TC Exposure Score of 1 by 4,442,607. To predict the number of cases of mental illness symptoms for Miami-Dade and Broward counties, the mental illness-specific predictions for the TC Sandy, 2004 FL TCs, and TC Ike were averaged.

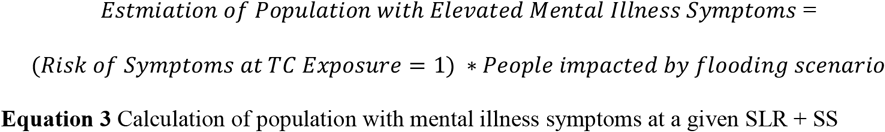

While a TC exposure score = 1 can confidently be related to the most extreme SLR + SS scenario, a TC exposure score = 1 may be likely from an even less severe scenario. Thus, we repeated these methods for 1.0 m SLR and a category 3 TC, a more likely future scenario given current efforts to mitigate the impacts of climate change. The population inundated for the later scenario was 4,208,923 people.

## RESULTS

### Regression Models

The logistic regressions showed several significant relationships between regressors and mental illness symptoms (OR 6). For the TC Sandy dataset, the resulting regression for post-TC MDD symptoms illustrated a positive relationship between higher risk of MDD symptoms and being Black (p = 0.004), having lifetime (pre-existing) anxiety (p<0.001), or experiencing greater TC exposure (p<0.001). Black subjects also showed lower risk of MDD symptoms as TC exposure increased (p = 0.033) (OR 7). Hispanic ethnicity (p=0.026), advanced age (p=0.003), lifetime (pre-existing) anxiety diagnoses (p = 0.021), lifetime (pre-existing) PTSD diagnoses (p = 0.003), or high TC exposure (p<0.001) were significantly associated with greater risk of PTSD symptoms. Greater risk of anxiety symptoms was positively associated with being female (p = 0.034), Hispanic (p = 0.03), having lifetime (pre-existing) anxiety (p<0.001), or having greater TC exposure (p<0.001).

For the TC Ike dataset, greater risk of MDD symptoms after TC exposure was positively related to being female (p = 0.004), having previous trauma exposure (p = 0.03), being Hispanic (p<0.001), and having lifetime MDD (p=0.018). Interestingly, better physical health (p = 0.046) and more informational support (p = 0.012) were also significantly correlated with greater risk of MDD symptoms. High risk of PTSD symptoms was positively correlated with being female (p = 0.001), Black (p = 0.034), or Hispanic (p = 0.003), having better health (p = 0.011), having lifetime PTSD (p = 0.005), and having higher TC exposure (p<0.001). Greater risk of anxiety symptoms was positively associated with having a lower income (p = 0.03), a higher health status (p = 0.006), a lower age (p = 0.001), or a lifetime anxiety diagnosis (p<0.001). There was also a significant interaction between income and TC exposure on elevated risk of anxiety symptoms (p = 0.011) (OR 7). In the setting of TC exposure, increased income level was positively associated with increased risk of anxiety symptoms after TC exposure.

The 2004 Florida TC dataset demonstrated that heightened risk of MDD symptoms post-TC was positively related to days displaced from one’s home (p = 0.015), higher health status (p<0.001), and lifetime PTSD diagnosis (p<0.001). Risk of PTSD symptoms was positively correlated to being male (p = 0.034). The risk of anxiety symptoms was related to having a lower income (p = 0.02), a higher health status (p = 0.028), lifetime MDD (p = 0.001), lifetime PTSD (p<0.001), or higher TC exposure (p<0.001). Similar to the Ike dataset, there was a significant interaction between income and TC exposure on anxiety symptoms (p = 0.029) (OR 7). Increasing income also increased the higher TC exposure’s positive correlation to greater risk of anxiety symptoms.

### TC Exposure

For a separate set of logistic regression models, only TC exposure was considered when determining elevated risk of mental illness symptoms (Fig. 3). Each of these models found that greater TC exposure was a significantly associated with developing mental illness symptoms. In the Florida subjects, TC exposure was positively correlated to risk of MDD (p<0.001), anxiety (p<0.001), and PTSD (p = 0.002) symptoms. The TC exposure score in the TC Ike subjects was positively associated with risk for heightened indications of MDD (p = 0.018), anxiety (p = 0.014), and PTSD (p<0.001) as well. These results were also similar among the TC Sandy subjects, with greater TC exposure associated with greater risk of MDD (p<0.001), anxiety (p<0.001), and PTSD (p<0.001) symptoms.

**Fig. 3.**
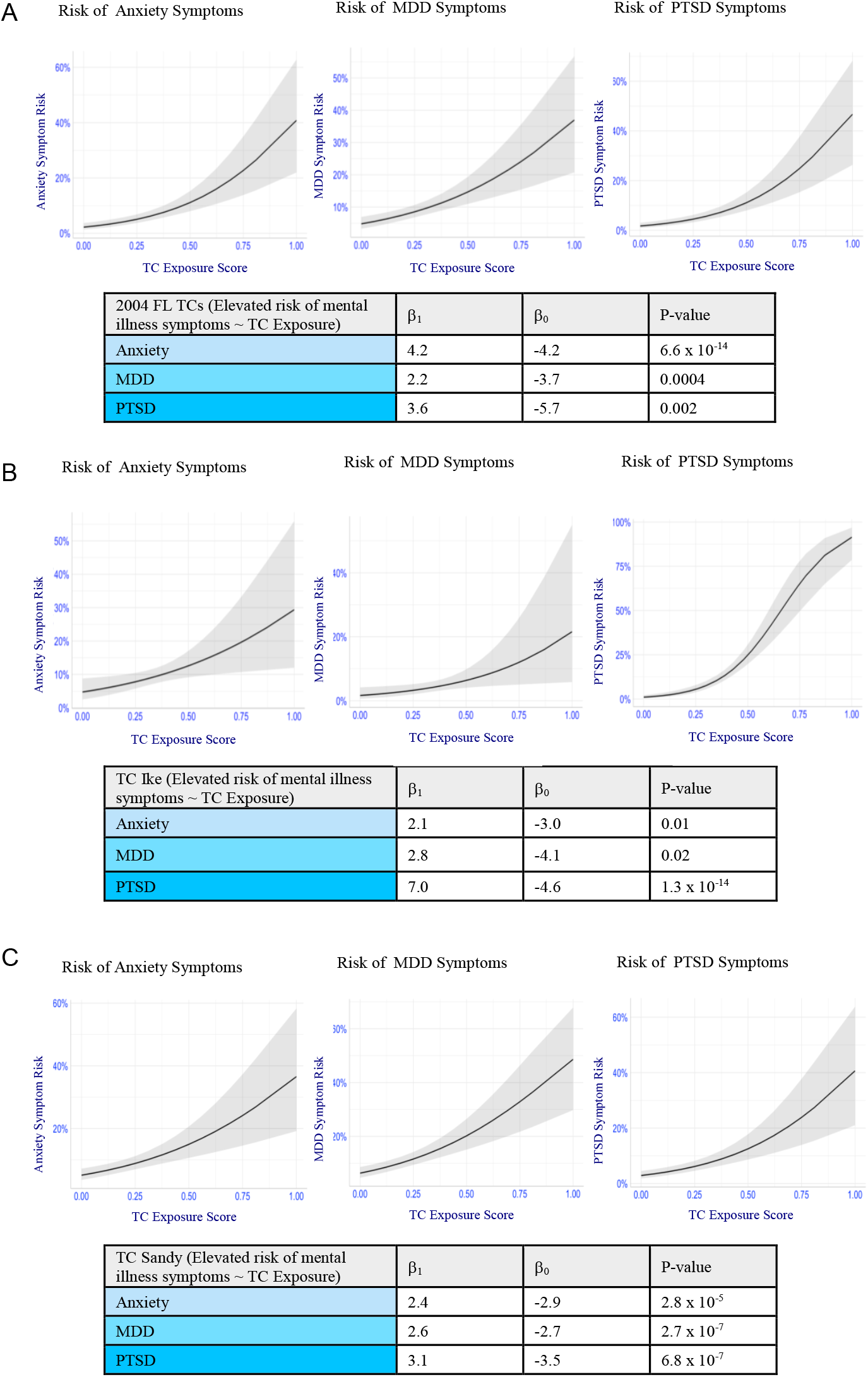
Graphs of mental illness risk vs tropical cyclone (TC) exposure and tropical cyclone exposure regression model results. Shaded area represents ±SEM. A) 2004 FL TCs B) 2008 TC Ike C) 2012 TC Sandy

When the sites were considered together via *meanz*, the TC exposure score was positively associated with risk of MDD (p<0.001), anxiety (p = 0.0014), or PTSD (p<0.001) symptoms (OR 8).

### Inundation Mapping

The inundation mapping of Miami-Dade and Broward counties showed significant flooding in all 29 scenarios (Fig. 4). Even the lowest predicted SLR, 0.5 meters, and no SS will impact over 180,000 people in the two counties. The dynamically mapped, lowest category TC SS scenario showed that SS will inundate more of the densely populated coast and less of the sparsely populated Florida Everglades region, impacting nearly 260,000 people. Combining the 2100 SLR estimates in the literature with predictions regarding TC frequency and intensity, a highly likely future scenario is 1-meter SLR and SS from a category 4 TC. Given this scenario, over 4 million people and 8,238 million m^2^ of land, almost the entirety of the two counties, would be devastated by inundation (Fig. 4). As the water level rises due to more extreme SLR and/or higher TC categories, almost the entirety of Miami-Dade and Broward counties is inundated, severely increasing the percentage of the population that is impacted (Fig. 5).

**Fig. 4.**
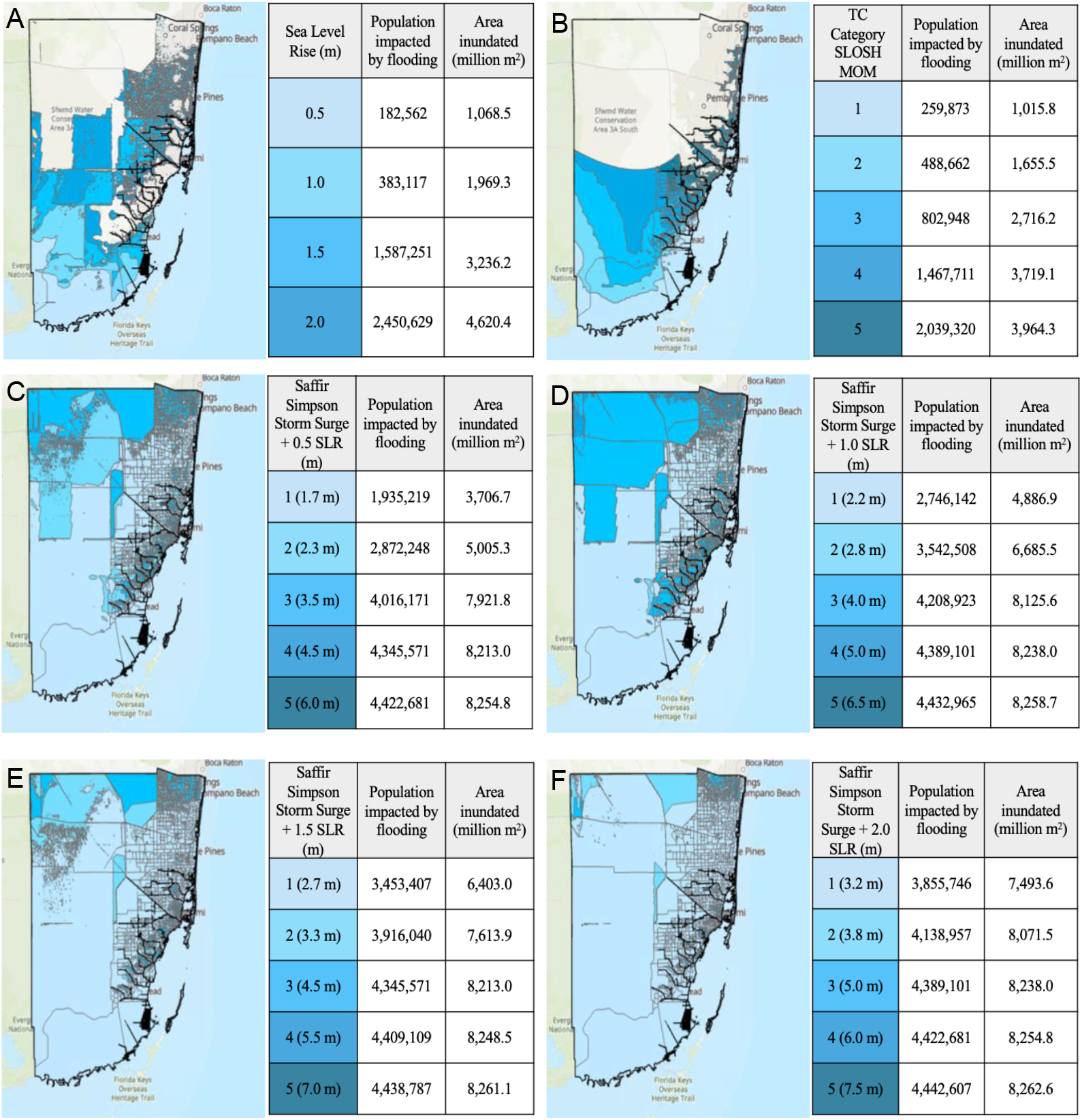
Inundation mapping of Miami-Dade and Broward counties for category 1 (light blue), category 2 (aqua), category 3 (bright blue), category 4 (teal), 5 (dark teal) storm surge and 0 through 2-meter sea level rise which are reported at 0.5-meter increments. The boxes represent the American Community Survey population boundaries A) Sea level rise only – 0.5 m (light blue), 1.0 m (aqua), 1.5 m (bright blue), 2.0 m (teal), B) Storm surge only C) 0.5 sea level rise + storm surge D) 1.0 sea level rise + storm surge E) 1.5 sea level rise + storm surge F) 2.0 sea level rise + storm surge

**Fig. 5.**
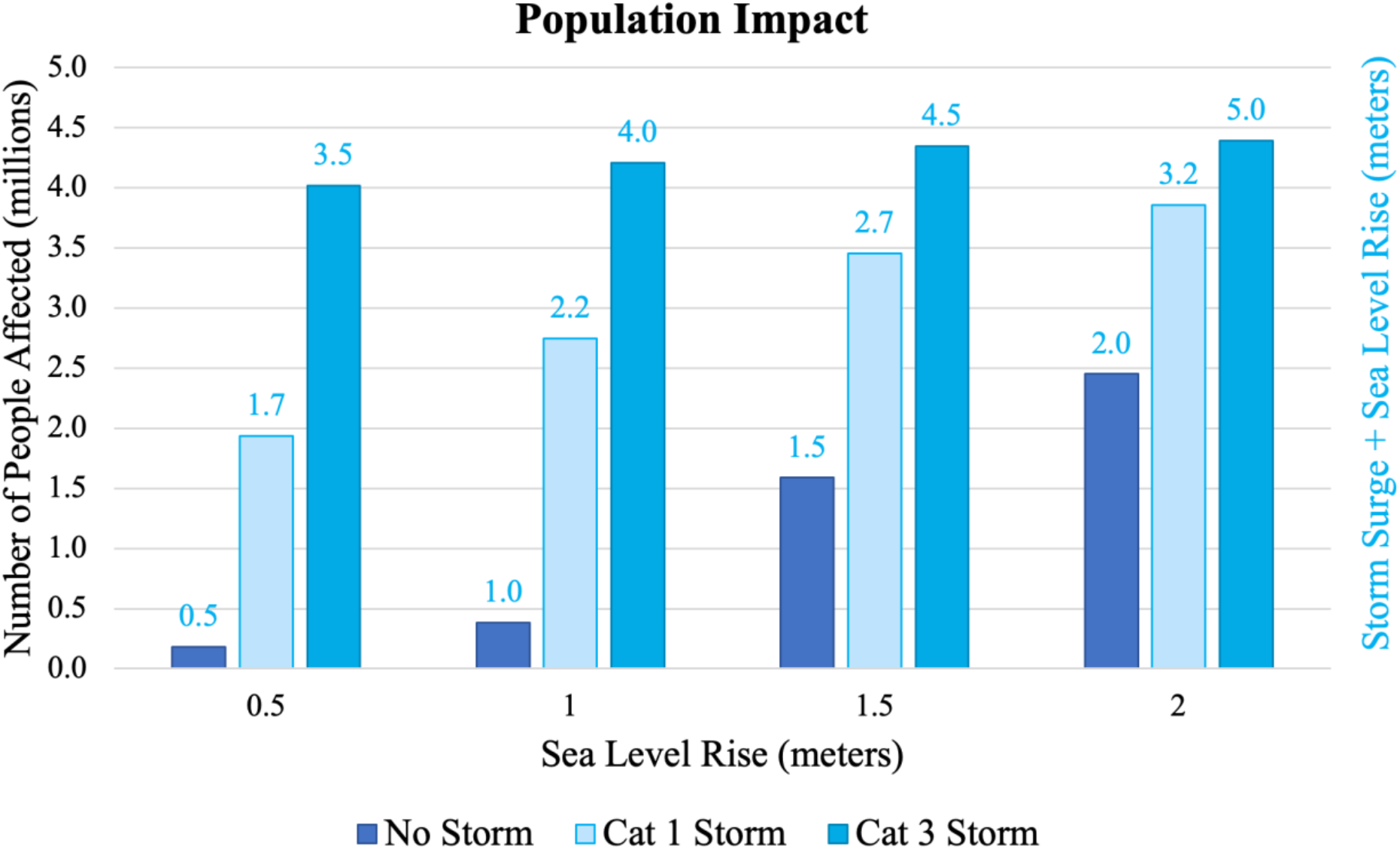
Illustration of the severe impacts of climate change induced natural disasters on Miami-Dade and Broward counties’ populations. The y-axis represents the population impacted by the scenarios, and the x-axis shows sea level rise. In each group (left to right 0.5 m, 1 m, 1.5 m, 2 m) leftmost bar is dark blue for no storm surge, middle bar is light blue for a category 1 storm (1.2 m SS), and rightmost bar is bright blue for a category 3 storm (3 m SS)

### Projected Mental Illness from SLR and TC Intensity Levels

We estimated the number of excess cases of anxiety, MDD, and PTSD symptoms in Miami Dade and Broward counties after TC exposure. Assuming that TC exposure = 1 corresponds to the worst-case scenario of 2.0 m SLR and a category 5 TC, we projected that 1,717,808 individuals would be have anxiety symptoms, 1,332,782 with MDD symptoms, and 2,102,833 with PTSD symptoms. Assuming a TC exposure score = 1 corresponds to a less extreme, but more likely scenario of 1.0 m SLR and a category 3 TC, we projected that 1,627,450 would be diagnosed with symptoms of anxiety, 1,262,676 with symptoms of MDD, and 1,992,223 with symptoms of PTSD (OR 9).

## DISCUSSION

This project aimed to demonstrate the profound impact of climate change-induced extreme weather events on the symptom prevalence of three common diagnoses of mental illness in South Florida. The results of regression modeling for three independent TC datasets, as well as the model combining results from these, strongly support that higher TC exposure is significantly related to a higher risk for symptoms of PTSD, anxiety, and MDD (Fig. 3; OR 8). While our results are concordant with the plethora of studies examining the prevalence of mental illness post-flooding and TC exposure (Acierno et al., 2007; An et al., 2019; Ferré et al., 2018; Galea et al., 2007; Ginexi et al., 2000; Jermacane et al., 2018; Kar, 2004; Lowe et al., 2013; McMillen et al., 2002; Nahar et al., 2014; Phifer et al., 1988; Pietrzak et al., 2012; Waelde et al., 2000; Waite et al., 2017), this study is unique in making *a priori* predictions on mental illness symptoms after TC exposure prior to the natural disaster, and equally importantly in linking realistic climate change-induced SLR and SS scenarios to the number of cases of mental illness symptoms (OR 9).

While TC exposure more broadly is positively correlated with mental illness symptom risk, we illustrate numerous other factors that may contribute to the elevated risk for symptoms of PTSD, anxiety, and MDD (OR 6). The regressors which frequently showed positive correlations to heightened risk of mental illness symptoms included Hispanic ethnicity, Black race, female sex, or a lifetime diagnosis (pre-existing) of mental health disorders.

There were also several variables which showed less consistent or unexpected associations with mental illness symptom risk. For instance, the inconsistent relationship between age and PTSD or MDD symptoms may be due to the wide range of age of onset for PTSD (median: 25-53 years; IQR: 15-75 years) and mood disorders (median: 25-45 years; IQR: 17-65 years) (Kessler, 2008). Another abnormal finding from the FL dataset showed that being male was positively correlated to a higher risk of PTSD symptoms. In all other instances where sex was significantly associated with heightened risk for symptoms of mental illness, however, females had a higher risk, in accordance with the literature (Essau et al., 2010; McLean, 2011). Thus, this discrepancy may be unique to the FL dataset. Also unique, the Sandy dataset showed that being Black lowered the risk of MDD symptoms as TC exposure increased (OR 7). The diverging relationships between age, sex, and race and risk of mental illness symptoms across the TC datasets, however, further demonstrate the impact that climate change will have on wide-ranging populations.

An intriguing relationship discovered from the regression models was a frequent positive correlation between better physical health and greater risk of mental illness symptoms. The actual prevalence of mental illness is difficult to determine given the many hurdles in accessing primary and mental healthcare (Adler and Ostrove, 1999). While multiple factors impact access to health care that were not explicitly considered, for the purpose of this project, these unexpected regression results may be due to a measurement error from lower income individuals having less access to healthcare, resulting in under-reporting of mental illness.

TC Ike data also showed an unexpected positive association between informational support and MDD symptom risk. Most studies suggest that post-disaster informational support reduces mental illness risk (Bryant et al., 2017; U.S. Department of Veterans Affairs, 2015). This abnormal result may be due to overwhelming or morbid information being shared.

The Florida and Ike datasets demonstrated that higher incomes increased the positive relationship between greater TC exposure and heightened risk of anxiety symptoms. This interaction was unexpected given the potential high costs of rectifying TC damage, paying for medical care, mitigating the financial burden of job loss, or reaching safety after a natural disaster. We may hypothesize that individuals with higher incomes may have more to lose, such as more property in locations more vulnerable to SS (e.g. beachfront), from extreme TCs. Higher income classes are more likely to have flood insurance, and more likely to report their TC exposures. Similarly, higher income groups will have better access to healthcare and are more likely to have diagnosed mental illness. Thus, these results may also be due to a measurement error.

While the logistic regression results clearly demonstrate the high risk of mental illness symptoms among individuals exposed to TCs, the inundation mapping stresses the widespread impact that climate change will have on society more broadly. Furthermore, given the immense number of people projected to develop anxiety, MDD, or PTSD symptoms in South Florida, we demonstrate the need for dedicating resources on a prodigious scale to addressing mental health as climate change threats intensify.

### Mitigation

To reduce the risk of SLR, TCs, and other climate change-induced consequences, carbon emissions must decrease. This goal could be achieved by improving public transportation accessibility, energy efficiency, and renewable energy availability (Hayes et al., 2018). Until efforts to reduce climate change are tangible, however, cities should plan for natural disasters by collecting supplies, deciding on evacuation versus sheltering (Espinel et al., 2019), and developing effective communication methods. While infrastructure and disaster plans can partially alleviate future mental health impacts, additional resources should be developed, such as community education on mental health resources, special training for emergency responders, identification of vulnerable populations, and research on climate change-induced mental health challenges (Bourque and Willox, 2014; Hayes et al., 2018).

Ultimately, a community’s adaptive capabilities are reliant on “governance, economics, infrastructure, technology, information and skills, institutions, and equity” (Séguin J, 2008). Thus, although mitigation of the impacts of climate changes may be possible for wealthy cities and countries, other threatened communities may be less resilient. Overall, dedicating more effort and resources into environmentally friendly initiatives would be an effective and accessible way to combat climate change and its catastrophic effects.

### Limitations

When studying mental health following TC exposures, it is vital to consider resilience, preparedness, and ability to adapt to climate change repercussions. The preparedness, resilience, and adaptability of a community depend on a number of factors, including community demographics, vulnerability, mental illness prevalence (Crimmins, 2016; Lowe et al., 2013; Tracy et al., 2011), and resource access. Additionally, many communities may migrate as a result of climate change induced natural disasters, further complicating our conclusions. In future studies, longitudinal data sources, rather than cross-sectional studies as used in this project, should measure community preparedness and adaptability. In this study, site-specific variations in preparedness and adaptability were not considered. Thus, the risks calculated for symptoms of PTSD, anxiety, and MDD may be under- or over-estimated.

Another limitation of this study was the lack of consideration of comorbidity. There are high levels of comorbidity between mood disorders, anxiety, and PTSD (Brown et al., 2001). Thus, by studying each of these illnesses individually, the risks may have been underestimated. Additionally, when considering pre-existing mental health conditions, which showed a positive association with risk of TC-related mental illness diagnosis, many individuals may not have had official diagnoses. Thus, the relationship between prior mental illness diagnosis and mental illness symptom risk may be over or under reported.

Missing data was imputed using the *mice* function. Imputation is imperfect, but multiple imputation using chained equations (White et al., 2011) can be problematic in datasets with numerous variables or with perfect prediction for binomial variables. Another potential problem with imputation is how similar the subjects with missing data are to the subjects whose data was used to impute values. Nevertheless, multiple imputation is deemed to be an effective method of handling missing data and was superior to removing subjects with missing data (Kang, 2013).

Finally, when conducting inundation mapping, static mapping was used for the SLR and SLR + SS scenarios. Static inundation mapping frequently overestimates flooding severity because it does not consider waves, landscape, sediment, or barriers (Vousdoukas, 2016). Regardless of these limitations, however, the extreme impact of climate change on mental health as demonstrated by this study are valid and incredibly important to consider.

### Strengths and Future Directions

This project expands on the scarce research relating mental health and climate change by assessing the risk of developing TC-associated mental illness symptoms including PTSD, MDD, and anxiety. Moreover, this project examines how various demographic characteristics, prior mental illness, post-disaster support can influence mental health after TC exposure using a sample of over 3,000 subjects. Future research should work to determine how various climate change-induced natural disasters, illnesses, and other consequences will impact community mental health in order to prepare accordingly.

The present study also elucidates the widespread impact that climate change-induced SLR and SS will have on high-risk communities. While previous studies have performed inundation mapping to exemplify the risk of SLR or SS, few have considered SLR and SS effects together. Future projects should aim to use dynamic inundation. Additionally, when using inundation data to predict mental illness symptom prevalence, researchers should collect TC exposure data directly related to flood levels in order to better predict how various inundation scenarios may impact the prevalence of mental illness symptoms.

## CONCLUSION

The results of the present study confirm the tremendous impact that TC exposure may have on mental illness symptom risk and demonstrates the profound number of people (millions) who may potentially be impacted by the climate change-induced stressors. Research has frequently focused on climate change in light of looming natural disasters, food shortages, polluted water, disease, and more, propelling many governments to establish protection plans to maintain infrastructure, economic strength, and the physical health of their communities. Despite the research that demonstrates that mental health will be greatly impacted by all of these stressors, insufficient research is focused directly on negative impact on mental health resulting from climate change. The present study underscores the need for expanded research on the intersection of climate change and mental health, and the importance of establishing mental health resources and protection plans before the devastation of climate change overwhelms vulnerable populations. Working as a global community, we can address the looming crises anticipated from climate change and the heightened risk of mental illness symptoms due to natural disasters.

## Data Availability

All datasets were approved use in this study under an exempt status granted by the Duke University Health System IRB. The tropical cyclone Sandy dataset was shared through a data use agreement (DUA: AGR-21494) between Mount Sinai Medical Center and Duke University Health System. The tropical cyclone Ike dataset was obtained through the Inter-University Consortium for Political and Social Research at University of Michigan. The 2004 FL tropical cyclones dataset was publicly available through contact with Dr. Ron Acierno. All subjects provided informed consent.
The USGS raster is available for download at www.usgs.gov. The ACS population data is available through ArcGIS as a Living Atlas feature layer from Esri.
Code availability: https://github.com/mmonsour567/More-than-an-increase-in-sea-levels-The- impact-of-climate-change-on-mental-illness-prevalence.git

https://github.com/mmonsour567/More-than-an-increase-in-sea-levels-The-impact-of-climate-change-on-mental-illness-prevalence.git

## Declarations

### Funding

This work was supported by the the Center for Disease Control and Prevention (CDC) (Grant number U01-TP000573-01).

### Conflicts of interest

Not applicable

### Availability of data and material

All datasets were approved use in this study under an exempt status granted by the Duke University Health System IRB. The tropical cyclone Sandy dataset was shared through a data use agreement (DUA: AGR-21494) between Mount Sinai Medical Center and Duke University Health System. The tropical cyclone Ike dataset was obtained through the Inter-University Consortium for Political and Social Research at University of Michigan. The 2004 FL tropical cyclones dataset was publicly available through contact with Dr. Ron Acierno. All subjects provided informed consent.

The USGS raster is available for download at www.usgs.gov. The ACS population data is available through ArcGIS as a Living Atlas feature layer from Esri.

## Acknowledgements

We acknowledge the contributions of Dr. Ron Acierno for sharing the 2004 Florida tropical cyclones dataset which was incredibly helpful for our analysis. We also acknowledge Dr. Sandro Galea and Dr. Fran Norris, who collected data for the Galveston Bay Recovery Study, obtained through the Inter-university Consortium for Political and Social Research.

Click here to access/download **Supplementary Material** online_resources.docx

## Notes

### Competing Interest Statement

The authors have declared no competing interest.

### Funding Statement

Dr. Emanuela Taioli's work was supported by the the Center for Disease Control and Prevention (CDC) (Grant number U01-TP000573-01). Dr. Taioli's dataset was used for data analysis in this work.

### Author Declarations

The DUHS IRB has determined that the following protocol meets the criteria for a declaration of exemption from further IRB review as described in 45 CFR 46.101(b), 45 CFR 46.102 (f), or 45 CFR 46.102 (d), satisfies the Privacy Rule as described in 45 CFR 164.512(i), and satisfies Food and Drug Administration regulations as described in 21 CFR 56.104, where applicable. Protocol ID: Pro00105402 Reference ID: Pro00105402-INIT-1.0 Protocol Title: The potential impact of rising sea levels and storm surge on mental health in Miami, FL Principal Investigator: Rajendra Morey Review Date: April 28, 2020 Expiration Date: *Does not expire Exempt Category: Category 4: Secondary research for which consent is not required: Secondary research uses of identifiable private information or identifiable biospecimens, if at least one of the following criteria is met: i. The identifiable private information or identifiable biospecimens are publicly available; ii. Information, which may include information about biospecimens, is recorded by the investigator in such a manner that the identity of the human subjects cannot readily be ascertained directly or through identifiers linked to the subjects, the investigator does not contact the subjects, and the investigator will not re-identify subjects; iii. The research involves only information collection and analysis involving the investigator's use of identifiable health information when that use is regulated under 45 CFR parts 160 and 164, subparts A and E [HIPAA], for the purposes of "health care operations" or "research" as those terms are defined at 45 CFR 164.501 or for "public health activities and purposes" as described under 45 CFR 164.512(b); or iv. The research is conducted by, or on behalf of, a Federal department or agency using government-generated or government-collected information obtained for nonresearch activities, if the research generates identifiable private information that is or will be maintained on information technology that is subject to and in compliance with section 208(b) of the E-Government Act of 2002, 44 U.S.C. 3501 note, if all of the identifiable private information collected, used, or generated as part of the activity will be maintained in systems of records subject to the Privacy Act of 1974, 5 U.S.C. 552a, and, if applicable, the information used in the research was collected subject to the Paperwork Reduction Act of 1995, 44 U.S.C. 3501 et seq. *This Declaration of Exemption from further IRB Review is in effect from April 28, 2020 and does not expire. However, changes to the proposed research will require an amendment requesting re-review for exemption. Reportable serious adverse events and unanticipated problems related to the research that place subjects or others at risk of physical, psychological, economic, or social harm must be promptly reported to the IRB and will result in reconsideration of the activity's exempt status.

