## Supplementary Information for "The Impact of Climate Change on the Prevalence of Mental Illness Symptoms"

Article title - The Impact of Climate Change on the Prevalence of Mental Illness Symptoms
Author names - Molly Monsour, Emily Clarke_Rubright, Wil Lieberman-Cribbin, Christopher Timmins, Emanuela Taioli, Rebecca M. Schwartz, Samantha S. Corley, Anna M. Laucis, Rajendra Morey
Affiliation of the corresponding author - Duke University

| Site | Sample Size | Males | Females | Black | Hispanic | Lifetime PTSD | Lifetime MDD | Lifetime Anxiety |
|---|---|---|---|---|---|---|---|---|
| 2004 Florida TCs | 1543 | 36% | 64% | 6.7% | 5.3% | 12.8% | 20% | NA |
| TC Ike | 658 | 40.1% | 59.9% | 15.7% | 19.5% | 12.2% | 22% | NA |
| TC Sandy | 818 | 29.6% | 71.4% | 18.1% | 16.7% | 3.3% | 12.6% | 11.2% |

**Online Resource 1** Site details

A

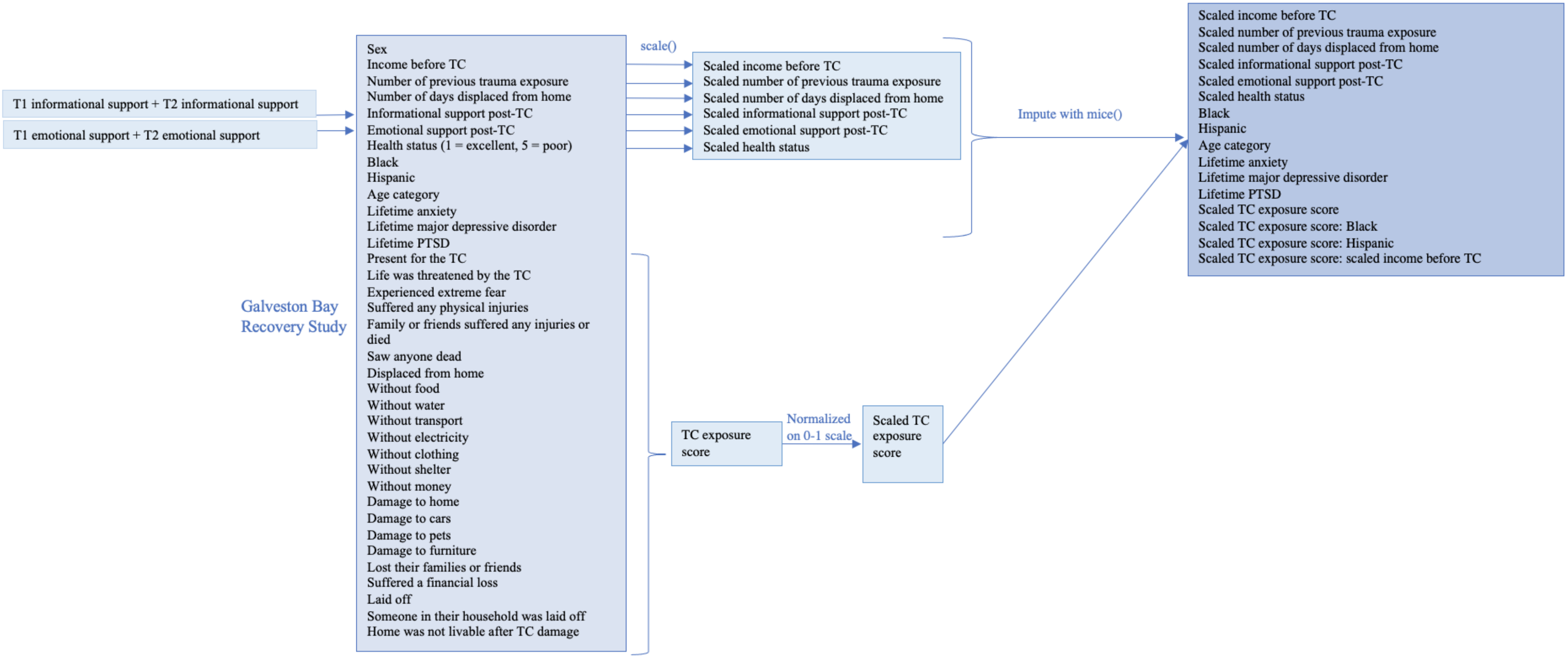

T1 informational support + T2 informational support
T1 emotional support + T2 emotional support
Galveston Bay Recovery Study
Sex
Income before TC
Number of previous trauma exposure
Number of days displaced from home
Informational support post-TC
Emotional support post-TC
Health status (1 = excellent, 5 = poor)
Black
Hispanic
Age category
Lifetime anxiety
Lifetime major depressive disorder
Lifetime PTSD
Present for the TC
Life was threatened by the TC
Experienced extreme fear
Suffered any physical injuries
Family or friends suffered any injuries or died
Saw anyone dead
Displaced from home
Without food
Without water
Without transport
Without electricity
Without clothing
Without shelter
Without money
Damage to home
Damage to cars
Damage to pets
Damage to furniture
Lost their families or friends
Suffered a financial loss
Laid off
Someone in their household was laid off
Home was not livable after TC damage
scale()
Scaled income before TC
Scaled number of previous trauma exposure
Scaled number of days displaced from home
Scaled informational support post-TC
Scaled emotional support post-TC
Scaled health status
TC exposure score
Normalized on 0-1 scale
Scaled TC exposure score
Impute with mice()
Scaled income before TC
Scaled number of previous trauma exposure
Scaled number of days displaced from home
Scaled informational support post-TC
Scaled emotional support post-TC
Scaled health status
Black
Hispanic
Age category
Lifetime anxiety
Lifetime major depressive disorder
Lifetime PTSD
Scaled TC exposure score
Scaled TC exposure score: Black
Scaled TC exposure score: Hispanic
Scaled TC exposure score: scaled income before TC

B

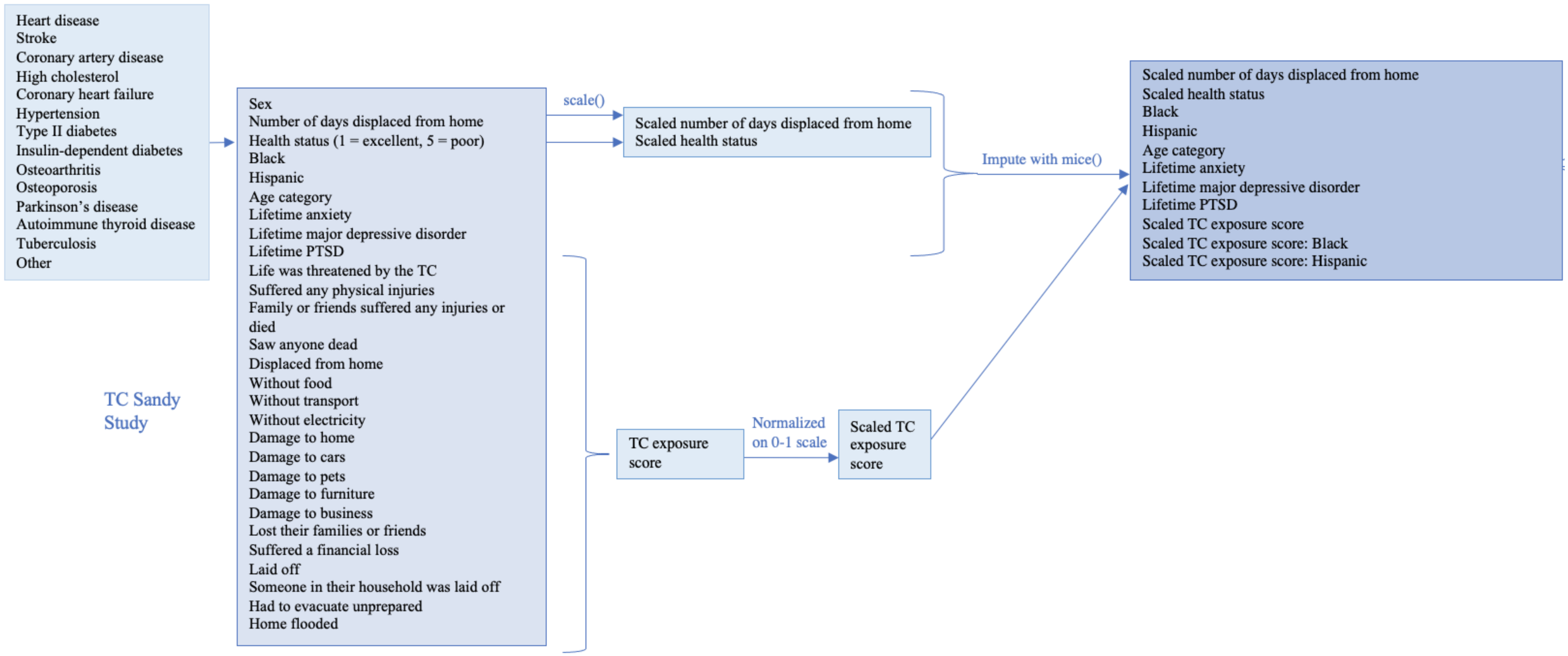

Heart disease
Stroke
Coronary artery disease
High cholesterol
Coronary heart failure
Hypertension
Type II diabetes
Insulin-dependent diabetes
Osteoarthritis
Osteoporosis
Parkinson's disease
Autoimmune thyroid disease
Tuberculosis
Other
TC Sandy Study
Sex
Number of days displaced from home
Health status (1 = excellent, 5 = poor)
Black
Hispanic
Age category
Lifetime anxiety
Lifetime major depressive disorder
Lifetime PTSD
Life was threatened by the TC
Suffered any physical injuries
Family or friends suffered any injuries or died
Saw anyone dead
Displaced from home
Without food
Without transport
Without electricity
Damage to home
Damage to cars
Damage to pets
Damage to furniture
Damage to business
Lost their families or friends
Suffered a financial loss
Laid off
Someone in their household was laid off
Had to evacuate unprepared
Home flooded
scale()
Scaled number of days displaced from home
Scaled health status
TC exposure score
Normalized on 0-1 scale
Scaled TC exposure score
Impute with mice()
Scaled number of days displaced from home
Scaled health status
Black
Hispanic
Age category
Lifetime anxiety
Lifetime major depressive disorder
Lifetime PTSD
Scaled TC exposure score
Scaled TC exposure score: Black
Scaled TC exposure score: Hispanic

C

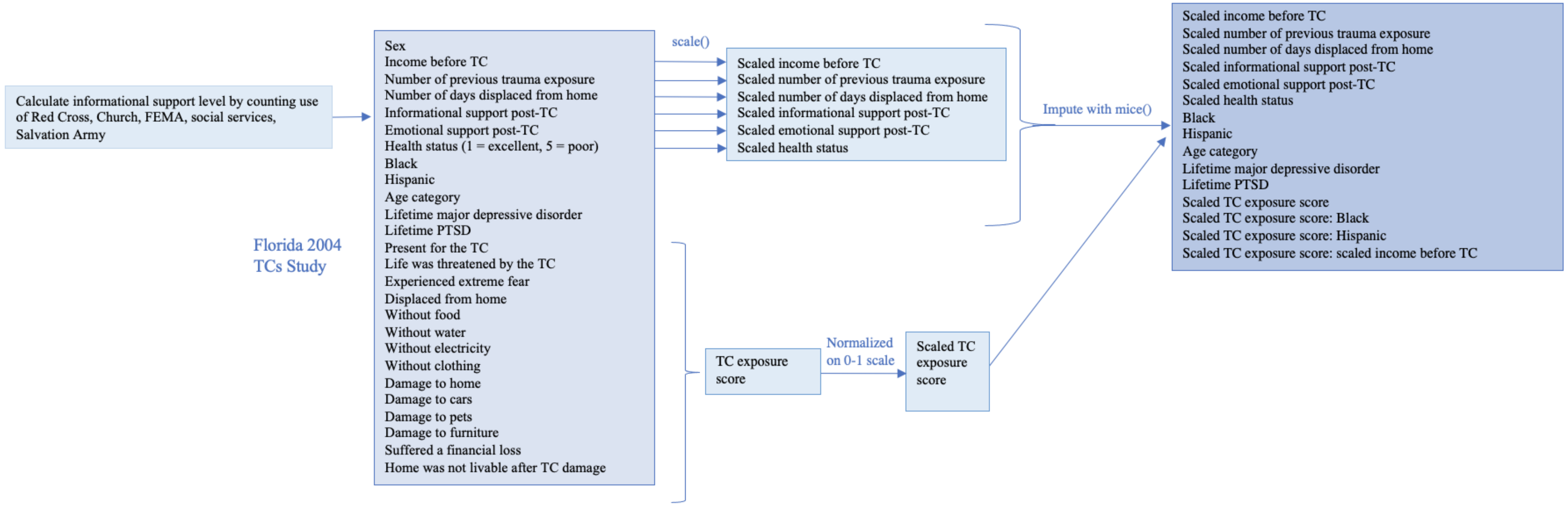


**Online Resource 2** Detailed description of independent variables considered for each site

A) TC Ike
B) 2004 FL TCs
C) TC Sandy

**Online Resource 3: Data Sites**

The first dataset used was the Galveston Bay Recovery Study (n=658), conducted by the National Center for Disaster Mental Health Research and obtained through the Inter-university Consortium for Political and Social Research (National Center for Disaster Mental Health Research et al., 2016). The data was collected by Galea and Norris. This data focused on the wellness of individuals impacted by the 2008 TC Ike. All subjects were living in Galveston or Chambers County before or during the TC. For the purpose of this study, the data collected at time point 2 (T2), 5-9 months after TC Ike, was used for analysis. Missing data was imputed by researchers who conducted the Galveston Bay Recovery Study; we used imputation version 5 as suggested in the dataset. For some survey questions asked at T2, the subjects were instructed to answer by reflecting on the time period after time period 1 (T1) and before T2. For these questions, we rescaled the T1 and T2 responses to reflect the subjects' experiences 5-9 months after the TC, rather than between the two time points. For example, if a subject rated informational support as a "3" 2-5 months after TC exposure, but received no informational support between T1 and T2, then the 3 and 0 ratings were added and rescaled to reflect the accurate informational support received 5-9 months after the TC.

The data collected from the 2004 Florida TCs was obtained from study team at The Medical University of South Carolina (Acierno et al., 2007). This study reflects the effects of four TCs (Charley, Frances, Ivan, and Jeanne), which demolished areas of Florida between August and September 2004. Unfortunately, this study did not include Miami-Dade or Broward counties due to the TCs having a greater impact along the Gulf of Mexico and Florida's west coast. The subjects were contacted through random digit dialing 6-9 months post TC, resulting in a sample size of 1,452 people. The data was weighted by gender and age to match the US Census 2000 data.

The study team at Mount Sinai Medical Center provided a dataset exploring the mental health impacts from TC Sandy (Schwartz et al., 2017). The subjects (n = 818) were from Nassau, Suffolk, Queens, and Richmond counties who were surveyed 11-28 months after the TC. Convenience sampling was used to recruit subjects, which may have produced biased results.

Various demographic measures were considered in the data analysis. Income level was divided into six categories for the Galveston dataset and eight for the Florida dataset. The Sandy dataset did not include income data. Six age groups were used in analysis, 18-24, 25-34, 35-44, 45-54, 55-64, and greater than 65. Race and ethnicity were also considered as binary variables for black race and/or Hispanic subjects. Health status was measured on a scale of 1-5, with 1 being Excellent, 2 being Very Good, 3 being Good, 4 being Fair and 5 being Poor. For the Sandy dataset, health status was not previously collected, but history of heart disease, stroke, coronary artery disease, high cholesterol, coronary heart failure, hypertension, type II diabetes, insulin-dependent diabetes, osteoarthritis, osteoporosis, Parkinson's disease, autoimmune thyroid disease, tuberculosis, and other were reported. To determine a health score, the incidence of each of these variables was added as +1 towards a composite health status score. A score of 0 translated to 1 for "excellent" health status, 1→2 (Very Good), 2→3 (Good), 3→4 (Fair), and >3→5 for Poor health status. Sex was coded as a binary variable (male = 1).

The two groups used different measures to identify elevated mental illness symptoms after TC exposure. To measure PTSD, the Florida subgroup used the National Women's Study PTSD Module (Kilpatrick et al. 1989), which was validated with the DSM-IV. The Galveston Study researchers used the PTSD Checklist-Civilian version and personalized questions in order to determine PTSD symptoms as a result of TC exposure (Ruggiero et al., 2003). Using a list of DSM-IV PTSD Criterion-A traumatic events , the Galveston Bay research group measured previous trauma of subjects. The Sandy study measured PTSD symptoms using used a TC Sandy-specific version of the Civilian PTSD Questionnaire (Ruggiero et al., 2003). Each study provided variables for lifetime PTSD and TC-linked PTSD symptoms, which were used in our analysis. The number of previous trauma exposures, reported in the Florida and Galveston data, were also used as an independent variable for these sites' analyses.

The 2004 Florida researchers measured major depressive disorder symptoms using the Structured Clinical Interview for DSM-IV (Spitzer RL, 1995). The Galveston study measured depressive symptoms using the Patient Health Questionnaire-9 (Kroenke K et al., 2001). Researchers at Mount Sinai used the Patient Health Questionnaire-4 (PHQ-4) to confirm MDD symptoms (Kroenke, 2009). The present project used the studies' reports of lifetime MDD and TC related MDD symptoms to analyze data.

To assess anxiety symptoms, the Florida study used the Structured Clinical Interview for DSM-IV (Spitzer RL, 1995). The Galveston research group used the General Anxiety Disorder-7 (GAD-7) (Kroenke, 2009) and additional questions. The Sandy dataset used the PHQ-4 to measure lifetime anxiety diagnoses and elevated anxiety symptoms after TC exposure. The Florida dataset did not report lifetime anxiety diagnoses; thus, lifetime anxiety was not considered as an independent variable for this site. Emergent anxiety symptoms after TC exposure were used as a dependent variable for all sites.

TC exposure was measured extensively by both groups. For analysis in this study, the various TC exposure variables measured by each group were added together to output a TC exposure score. The Galveston group's exposure score was determined based on whether the subject was present for the TC, life was threatened from the TC, experienced extreme fear, suffered any physical injuries, had family or friends who suffered any injuries or died, witnessed death, was displaced from home, was without food, water, transport, electricity, clothing, shelter, or money, whether home, cars, pets, or furniture were damaged, lost family members or friends, suffered a financial loss, was laid off, household member was laid off, or had an unlivable home after TC damage. The Florida group's scores were measured by whether the subject was present for the TC, life was threatened from the TC, experienced extreme fear, was displaced from home, was without food, electricity, water, or clothing, whether home, cars, pets, or furniture were damaged, suffered a financial loss, or had an unlivable home after TC damage. The Sandy group TC exposure score was built from  whether the subject's life was threatened from the TC, suffered any physical injuries, family or friends suffered any injuries or died, saw anyone dead, was displaced from home, without food, transport, or electricity,  whether home, cars, pets, or furniture were damaged, lost their families or friends, suffered a financial loss, was laid off, someone in their household was laid off,  had to evacuate unprepared, businesses were damaged, or home was flooded. Once a composite score was calculated, each subject was normalized to a score between 0 and 1 via the minimum and maximum scores of the subject's site.

$$TC\ Exposure\ Score = \frac{TC\ Exposure\ Count - minimum(TC\ Exposure\ Count)}{maximum(TC\ Exposure\ Count)}$$

Due to the impact that informational and social support can have on mental illness (Kaniasty, 2012), social support and informational support post-TC were also analyzed. For the Florida dataset, I coded informational support by calculating the number of informational resources (i.e. Red Cross, Church, FEMA, social services, Salvation Army) available to each participant. Based on the number of resources, I ranked the level of informational support as low (0-2 resources), medium (3-4 resources), or high (5-6 resources). The social support variable in the Florida dataset was split into three categories, with 1 representing low support and 3 representing high support. Three aspects of social support were measured in the study: emotional, instrumental, and appraisal. Based on a 4-point scale, subjects answered questions related to these aspects of social support. The scores were divided into low (lowest third), medium, and high (highest third) (National Center for Disaster Mental Health Research et al., 2016). For the Galveston study, informational support and emotional support were measured via the Inventory of Post-disaster Social Support (Kaniasty, 2012). Galveston variables were recoded to include all time before T2, rather than the time between T1 and T2. The Sandy dataset did not supply measures of social or informational support.

<u>Online Resource 3 References</u>

Acierno, R., Ruggiero, K.J., Galea, S., Resnick, H.S., Koenen, K., Roitzsch, J., de Arellano, M., Boyle, J., Kilpatrick, D.G., 2007. Psychological sequelae resulting from the 2004 Florida hurricanes: implications for postdisaster intervention. Am J Public Health 97 Suppl 1, S103-108.

Kaniasty, K.Z., 2012. Predicting social psychological well-being following trauma: The role of postdisaster social support. Psychological Trauma: Theory, Research, Practice, and Policy 4, 22-33.

Kroenke K, Spitzer R L, B, W.J., 2001. The PHQ-9: validity of a brief depression severity measure. Journal of General Internal Medicine 16, 606-613.

Kroenke, K., Spitzer, R. L., Williams, J. B., & Lowe, B. , 2009. An Ultra-Brief Screening Scale for Anxiety and Depression: The PHQ–4. Psychosomaticss 50, 613-621.

National Center for Disaster Mental Health Research, Galea, S., Norris, F., 2016. Galveston Bay Recovery Study, 2008-2010, Ann Arbor, MI: Inter-university Consortium for Political and Social Research.

Ruggiero, K.J., Del Ben, K., Scotti, J.R., Rabalais, A.E., 2003. Psychometric properties of the PTSD checklist-civilian version. J Trauma Stress 16, 495-502.

Schwartz, R.M., Gillezeau, C.N., Liu, B., Lieberman-Cribbin, W., Taioli, E., 2017. Longitudinal Impact of Hurricane Sandy Exposure on Mental Health Symptoms. Int. J. Environ. Res. Public Health 14, 1-12.

Spitzer RL, W.J., Gibbon M, et al., 1995. Structured Clinical Interview for DSM-IV. . American Psychiatric Press, Washington, DC.

**Online Resource 4: Calculation of population impact**

The population data for Miami-Dade and Broward counties came from the ArcGIS layer, ACS Population Variables – Boundaries (Online Resource 5). This feature layer includes many demographic variables from the most recent American Community Survey (2019) (U.S. Census Bureau, 2019). The boundaries used for the counties are based on the US Census TIGER geodatabases. For this project, we focused primarily on the land area of each tract, the water area of each tract, and the total population per tract.

$$\frac{Area\ of\ Polygon\ Inundated}{Total\ Land\ Area\ of\ Polygon + Total\ Water\ Area\ of\ Polygon} \times Total\ Population$$

**Equation 2** Calculation of population impacted by inundation scenarios

This method poses a potential limitation by assuming equal distribution of the population, as some areas of larger polygons may be more population dense than others.

Online Resource 4 References

U.S. Census Bureau, 2019. American Community Survey 5-Year Data (2009-2018).

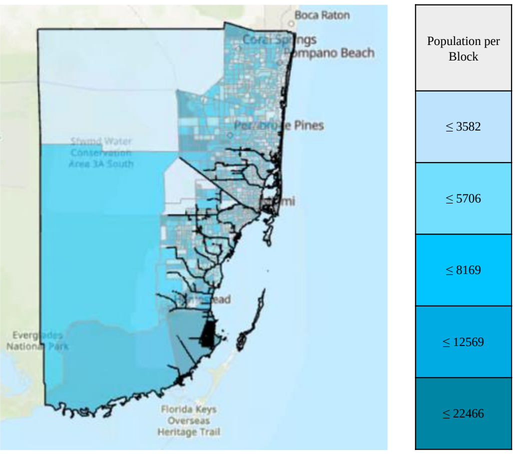


**Online Resource 5** Miami Dade and Broward  counties population mapped by block.

| Florida (MDD symptoms after TC Exposure~) | β value | Standard Error | P-value |
|---|---|---|---|
| **Days Displaced** | 0.220 | 0.090 | **0.015*** |
| **Health Status** | 0.507 | 0.139 | **0.0003***** |
| **Age Category** | -0.247 | 0.147 | **0.093*** |
| Lifetime MDD | 19.629 | 811.683 | 0.981 |
| **Lifetime PTSD** | 0.999 | 0.291 | **0.0006***** |

| Florida (PTSD symptoms after TC Exposure~) | β value | Standard Error | P-value |
|---|---|---|---|
| **Male** | 1.175 | 0.555 | **0.034*** |
| Income | -0.445 | 0.263 | 0.09 |
| Hispanic | -4.196 | 3.267 | 0.199 |
| Lifetime MDD | 1.144 | 0.808 | 0.157 |
| Lifetime PTSD | 19.383 | 1257.077 | 0.988 |
| TC Exposure | 1.585 | 1.383 | 0.252 |
| Hispanic: TC Exposure | 8.001 | 6.130 | 0.192 |

| Florida (Anxiety symptoms after TC Exposure~) | β value | Standard Error | P-value |
|---|---|---|---|
| **Income** | -0.624 | 0.268 | **0.020*** |
| **Health Status** | 0.264 | 0.120 | **0.028*** |
| Hispanic | 0.590 | 0.381 | 0.121 |
| **Lifetime MDD** | 0.983 | 0.291 | **0.001***** |
| **Lifetime PTSD** | 1.469 | 0.286 | **$2.8 \times 10^{-7}$***** |
| **TC Exposure** | 3.791 | 0.633 | **$2.09 \times 10^{-9}$***** |
| **Income: TC Exposure** | 1.371 | 0.629 | **0.029*** |

| Ike (MDD symptoms after TC Exposure~) | β value | Standard Error | P-value |
|---|---|---|---|
| **Male** | -1.879 | 0.649 | **0.004**** |
| Income | 0.347 | 0.244 | 0.155 |
| **Informational Support** | 0.570 | 0.227 | **0.012*** |
| Days Displaced | -0.586 | 0.384 | 0.127 |
| **Number of Previous Trauma** | 0.470 | 0.217 | **0.030*** |
| **Health Status** | 0.447 | 0.224 | **0.046*** |
| **Hispanic** | 1.933 | 0.477 | **$5.02 \times 10^{-5}$***** |
| **Lifetime MDD** | 1.076 | 0.453 | **0.018*** |

| Ike (PTSD symptoms after TC Exposure~) | β value | Standard Error | P-value |
|---|---|---|---|
| **Male** | -1.112 | 0.344 | **0.001***** |
| Income | 0.236 | 0.158 | 0.135 |
| Informational Support | 0.284 | 0.146 | 0.052 |
| **Health Status** | 0.386 | 0.152 | **0.011*** |
| **Black** | 0.795 | 0.375 | **0.034*** |
| **Hispanic** | 1.032 | 0.347 | **0.003**** |
| Age Category | 0.177 | 0.097 | 0.069 |
| Lifetime Anxiety | 0.719 | 0.384 | 0.061 |
| **Lifetime PTSD** | 1.003 | 0.359 | **0.005**** |
| **TC Exposure** | 6.659 | 1.054 | **$2.7 \times 10^{-10}$***** |

| Ike (Anxiety symptoms after TC Exposure~) | β value | Standard Error | P-value |
|---|---|---|---|
| Male | -0.692 | 0.398 | 0.082 |
| **Income** | -1.093 | 0.504 | **0.030*** |
| **Health Status** | 0.536 | 0.196 | **0.006**** |
| **Age Category** | -0.417 | 0.122 | **0.001***** |
| **Lifetime Anxiety** | 3.911 | 0.379 | **$<2 \times 10^{-16}$***** |
| TC Exposure | 0.807 | 1.276 | 0.527 |
| **Income: TC Exposure** | 3.438 | 1.344 | **0.011*** |

| Sandy (MDD symptoms after TC Exposure~) | β value | Standard Error | P-value |
|---|---|---|---|
| **Black** | 1.118 | 0.389 | **0.004**** |
| **Lifetime Anxiety** | 1.437 | 0.275 | **$1.86 \times 10^{-7}$***** |
| **TC Exposure** | 3.334 | 0.596 | **$2.24 \times 10^{-8}$***** |
| **Black: TC Exposure** | -3.175 | 1.490 | **0.033*** |

| Sandy (PTSD symptoms after TC Exposure ~) | β value | Standard Error | P-value |
|---|---|---|---|
| **Hispanic** | 0.832 | 0.375 | **0.026*** |
| **Age Category** | 0.299 | 0.101 | **0.003**** |
| **Lifetime Anxiety** | 0.907 | 0.393 | **0.021*** |
| **Lifetime PTSD** | 1.548 | 0.513 | **0.003**** |
| **TC Exposure** | 2.990 | 0.671 | **$8.38 \times 10^{-6}$***** |

| Sandy (Anxiety symptoms after TC Exposure ~) | β value | Standard Error | P-value |
|---|---|---|---|
| **Male** | -0.759 | 0.357 | **0.034*** |
| **Hispanic** | 0.691 | 0.318 | **0.030*** |
| **Lifetime Anxiety** | 2.213 | 0.290 | **$2.14 \times 10^{-14}$***** |
| **TC Exposure** | 2.319 | 0.627 | **$2.17 \times 10^{-4}$***** |

**Online Resource 6** Full logistic regression model results (TC = tropical cyclone)

A

**Income: TC Exposure Interaction for Anxiety Symptoms After 2004 FL TCs**

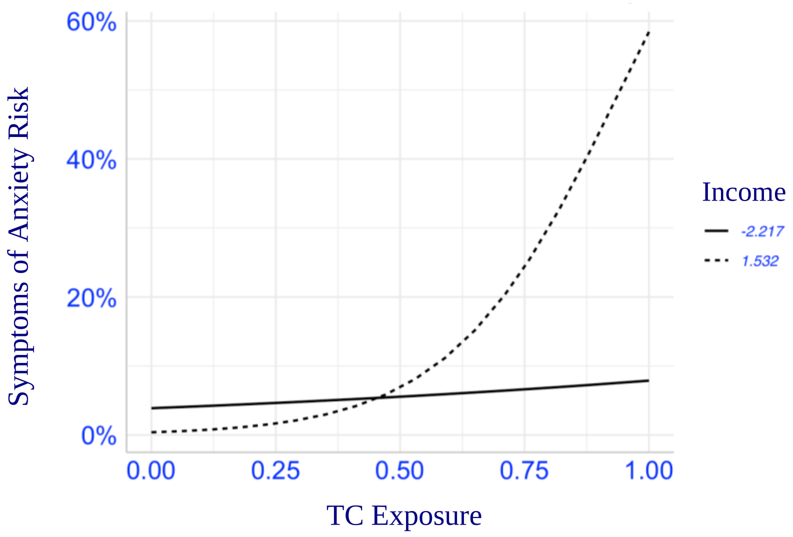


**Income: TC Exposure Interaction for Anxiety Symptoms After TC Ike**

B

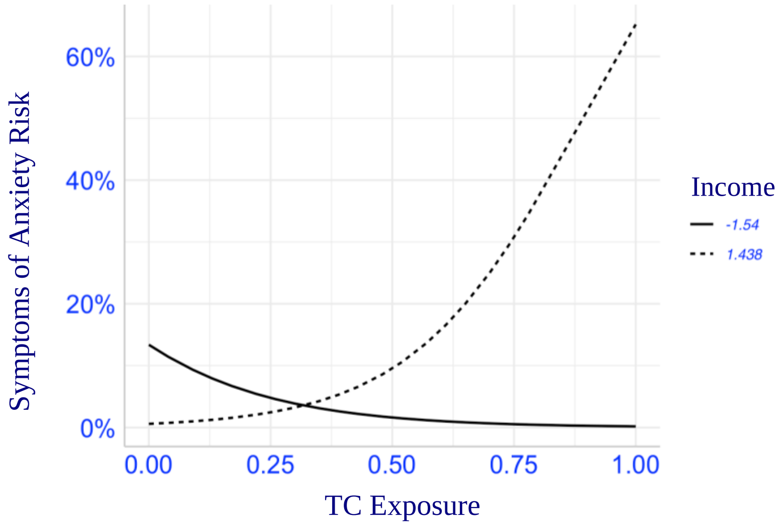


**Black: TC Exposure Interaction for MDD Symptoms After TC Sandy**

C

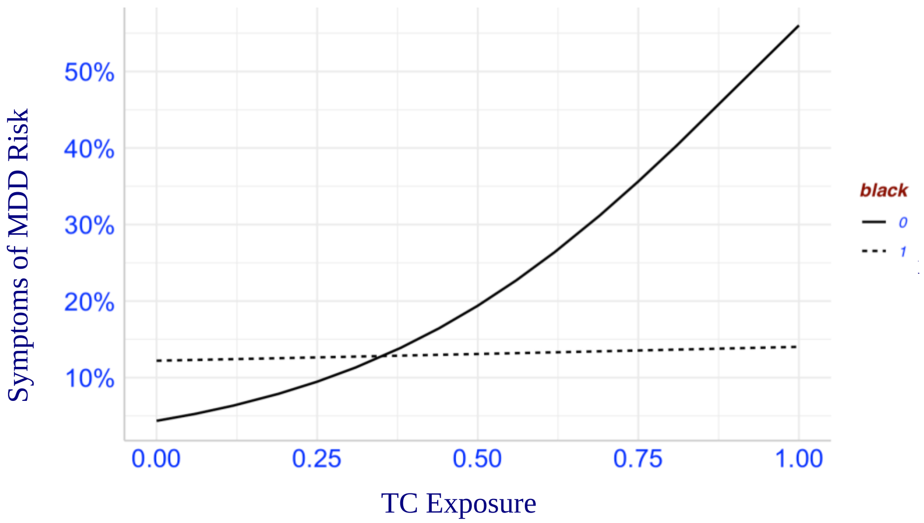

**Online Resource 7** The influence of maximum and minimum scaled income levels on anxiety symptoms after TC exposure if the interaction was significant for the site and the influence of Black race on MDD after TC Sandy

A) Income: TC Exposure for Anxiety Symptoms after 2004 FL TCs
B) Income: TC Exposure for Anxiety Symptoms after TC Ike
C) Black: TC Exposure for MDDD Symptoms after TC Sandy

| Elevated Mental Illness Symptoms | meanz() | p-value |
|---|---|---|
| Generalized Anxiety Disorder | 2.99 | 0.0014 |
| Major Depressive Disorder | 4.33 | $7.59 \times 10^{-6}$ |
| PTSD | 3.66 | $1.26 \times 10^{-4}$ |

**Online Resource 8** TC exposure regression model results after *meanz*()

| | Predicted Risk if TC Exposure Score = 1 | Additional reported elevated symptoms of mental illness cases = predicted risk X population impacted (TC Exposure = 1 is SLR = 1.0 m and TC Category = 3) | Additional reported elevated symptoms of mental illness cases = predicted risk X population impacted (TC Exposure = 1 is SLR = 2.0 m and TC Category = 5) |
|---|---|---|---|
| TC Sandy Anxiety Symptoms | 0.37 | 1,557,301 | 1,643,764 |
| TC Sandy MDD Symptoms | 0.49 | 2,062,372 | 2,176,877 |
| TC Sandy PTSD Symptoms | 0.41 | 1,725,658 | 1,821,468 |
| TC Ike Anxiety Symptoms | 0.29 | 1,220,587 | 1,288,356 |
| TC Ike MDD Symptoms | 0.22 | 925,963 | 977,373 |
| TC Ike PTSD Symptoms | 0.91 | 3,830,119 | 4,042,772 |
| 2004 FL TCs Anxiety Symptoms | 0.5 | 2,104,461 | 2,221,303 |
| 2004 FL TCs MDD Symptoms | 0.19 | 799,695 | 844,095 |
| 2004 FL TCs PTSD Symptoms | 0.1 | 420,892 | 444,260 |
| Average Anxiety Symptoms | --- | 1,627,450 | 1,717,808 |
| Average MDD Symptoms | --- | 1,262,676 | 1,332,782 |
| Average PTSD Symptoms | --- | 1,992,223 | 2,102,833 |

**Online Resource 9** Estimates for Miami Dade and Broward counties elevated symptoms of mental illnesses based on population impacted by inundation scenarios and logistic regression models (mental illness symptom risk ~ TC Exposure Score).
